# Long COVID Longitudinal Symptoms Burden Clusters Within A National Community-Based Cohort

**DOI:** 10.1101/2025.11.01.25339283

**Authors:** Yanhan Shen, Zach Shahn, McKaylee M. Robertson, Kelly Gebo, Denis Nash, the CHASING COVID Cohort Study Team

## Abstract

**Background:** Long COVID is clinically heterogeneous, with evolving symptom trajectories that complicate classification. Prior clustering studies often rely on Electronic Health Records data, risking underreporting. We used longitudinal, community-based data to characterize symptom clusters and risk factors.

**Methods:** We analyzed CHASING COVID Cohort participants with confirmed SARS-CoV-2 infection between December 2020 and December 2022, ≥12 months of follow-up, and long COVID (≥1 new symptom and concurrent activity limitation 3–12 months post-infection, both absent pre-infection). The infection in this window was the index infection; those with pre-index long COVID were excluded. Symptoms were self-reported pre-infection and at ∼3, 6, 9, and 12 months. Missing data were handled via multiple imputation by chained equations (30 datasets). Longitudinal K-means clustering was performed within each imputed dataset, with hierarchical aggregation to derive final assignments. Multivariable logistic regression (adjusting for age, sex) assessed associations of demographic, clinical, and social factors with cluster membership. Within the highest-burden cluster, hierarchical clustering identified symptom phenotypes.

**Results:** Of 1,787 infected participants, 511 met criteria (22% ≥50 years; 55% female; 60% White non-Hispanic; 54% mental health disorder; 7.2% immunodeficiency; 21% ≥2 comorbidities; 30% prior infection; 24% never vaccinated pre-index). Three clusters emerged: highest, moderate, and lowest burden. The highest-burden cluster (median 6 symptoms at 6 and 9 months) was characterized by fatigue (57%–74%), concentration difficulty, post-exertional malaise, myalgia, sleep disturbance, gastrointestinal symptoms, headache, irritability, and mobility limitations (each ∼46%–53%). The moderate cluster peaked at 3 symptoms (fatigue 42%); the lowest remained ∼1 symptom (fatigue 24% at 12 months). Older age (aOR 2.68, 95% CI 1.59–4.53), female sex (2.36, 1.48–3.76), mental health disorder (2.65, 1.65–4.25), and immunodeficiency (4.23, 1.71–10.51) were associated with highest vs lowest burden. Within the highest-burden cluster, three phenotypes emerged: neurological/multisystemic, psychiatric/neurological, and physical/respiratory.

**Conclusions:** Distinct long-COVID clusters and phenotypes underscore heterogeneity and support tailored management and risk stratification.

## INTRODUCTION

Long COVID is a complex condition characterized by substantial breadth, with diverse symptoms—including fatigue, respiratory, neurological, cardiovascular, and gastrointestinal manifestations—spanning multiple organ systems ^1,2^. Its substantial depth arises from complex underlying mechanisms such as viral persistence, immune dysregulation, endothelial dysfunction, autonomic disturbances, and mitochondrial impairment, presenting significant clinical and research challenges ^1,3,4^. Clustering analysis offers a valuable tool to uncover symptom heterogeneity and enhance our understanding of disease pathophysiology ^5–13^.

Clustering algorithms analyze data distribution and patterns to group patients with similar characteristics ^5,14,15^. Longitudinal studies utilizing electronic health records (EHR) data collected at multiple time points have identified distinct phenotypes of long COVID, characterized by varying symptom clusters and trajectories across different stages of recovery. For instance, a two-year prospective study observed that long COVID symptoms often peak between 6 to 12 months post-infection, significantly impacting the quality of life during this period ^16^. Another longitudinal study employing latent class analysis on patient-reported outcomes across multiple time points identified distinct symptom profiles, highlighting the heterogeneous nature of symptom presentation over time ^8^. Additionally, a study clustering participants based on symptoms documented at both 9 and 12 months post-infection identified four clusters: no or minor symptoms; multi-symptoms; joint pain; and neurocognitive-related symptoms, which were similar at both 9 and 12 months post-infection ^17^. Collectively, these findings emphasized that symptom clustering in long COVID is dynamic and changes over time and underscored the importance of continuous monitoring and personalized management strategies for individuals experiencing long COVID.

There are limitations to using EHR data to cluster long COVID patients’ symptoms due to under-reporting or under-ascertainment of milder or transient symptoms and the inability to adequately capture subjective experiences such as disabilities and limitations in daily life ^6,8–12,18^. Furthermore, since EHR data rely on individuals actively engaging in healthcare, they often fail to capture longitudinal long COVID symptoms ^19^. In contrast to EHR-based studies, which are limited to individuals actively engaging with the healthcare system and often miss milder or subjective symptoms, community-based cohort studies include individuals both within and outside of the healthcare system and can capture longitudinal, self-reported symptoms, providing a more comprehensive picture of long COVID’s progression and impact over time.

We aimed to leverage the community-based data to identify symptom clusters of participants with long COVID based on systematically ascertained self-reported symptoms. We also aimed to identify demographic, socioeconomic, and comorbidity characteristics of those experiencing the highest symptom burden.

## METHODS

### Participants

The Communities, Households, and SARS-CoV-2 Epidemiology (CHASING) COVID Cohort study was a national prospective cohort, which recruited participants between March 28, 2020 and August 21, 2020 ^20^. The community-based sample included adults residing in 50 U.S. states, the District of Columbia, Puerto Rico, and Guam. Study participants were recruited through social media ads (Facebook, Instagram, Scruff), Qualtrics Panel, and referrals from personal networks. Advertisements were written in English and Spanish, targeting people in the U.S. and U.S. territories aged 18 and older. As part of the study design, participants completed approximately quarterly surveys on COVID-19 exposures, symptoms, and testing, and also completed repeat serologic testing for SARS-CoV-2 infection approximately annually. Details of the study design, recruitment procedures, and serology-based incidence in this cohort are described elsewhere ^21–23^. The study protocol was approved by the Institutional Review Board of the City University Of New York (CUNY) (New York, NY, USA). Participant consent was obtained online at baseline and periodically during follow-up assessments.

### Pre- and Post-infection Symptoms and Activity Limitations

In each follow-up assessment since November 2020 approximately every three months, participants were asked whether they were currently experiencing any of the following symptoms: shortness of breath, difficulty walking more than 15 minutes, difficulty running/exercising, fatigue, post-exertional malaise, headache, trouble concentrating/brain fog, dizziness, irritability, erratic heartbeat, gastrointestinal issues, low-grade fever, myalgia, loss or alteration of taste/smell, and difficulty sleeping. We categorized the longitudinal symptom assessments into pre-infection, 3-, 6-, 9-, and 12-month post-infection periods based on the number of days between the assessment and the infection dates (determination of SARS-CoV-2 infection status and dates is outlined in **Appendix 1**). Specifically, assessments conducted 45-365 days before infection were classified as pre-infection data, while those within 45-135, 136-225, 226-315, and 316-405 days post-infection were categorized as 3-, 6-, 9- and 12-month post-infection, respectively. If participants completed more than one symptom assessment within a given interval, only the response closest to the target time point (i.e., 90, 180, 270, or 360 days post-infection) was retained to avoid potential overcounting of symptoms.

To address missing symptoms data, we applied multiple imputation by chained equation (MICE) with the fully conditional specification (FCS), generating 30 imputed datasets ^24,25^. Given the non-monotone missing data pattern and the assumption of missing at random, we accounted for occasional missed follow-ups and timing constraints that prevented appropriate interval assignment. We assumed that missing symptom data resulted either from participants occasionally missing follow-up assessments or from the timing of the follow-up assessment fielding, which sometimes prevented us from assigning the appropriate follow-up intervals. Variables selected for the imputation model included those correlated with the missing symptom data and predictive of missingness in follow-ups ^26^. A logistic regression model was used for each outcome, given the outcomes at other time points and the covariates ^27^. Specifically, we included the following variables as the dependent variables in the logistic regression model: symptom data from prior time points, age, gender, race/ethnicity, education, number of comorbidities, employment category, geographic region of residence, and material hardship. Details on demographics, socioeconomic factors, comorbidities, prior infection history, and COVID-19 vaccination status are provided in **Appendix 2**.

Participants were classified as having activity limitations if they reported difficulty in daily activities or household responsibilities ^28^. At each follow-up, participants who reported “Some difficulty” or “A lot of difficulty” to a question assessing challenges in engaging in daily activities or household responsibilities due to physical, mental, or emotional problems—one of the two-item disability measurement from the BRFSS survey—were considered to have activity limitations ^29^.

### Long COVID Case Definition

We defined long COVID cases as participants who experienced at least one symptom once between 3 and 12 months post-infection that was not present in the year preceding the index infection date, in combination with concurrent activity limitations which was present at the same time of symptom and absent prior to the index infection. The long COVID case definition was informed by guidance from the World Health organization (WHO) ^30^, the Centers for Disease Control and Prevention (CDC) ^31^, and the National Academies of Science, Engineering, and Medicine (NASEM) ^32^, all of which emphasize new or ongoing symptoms and functional impairment occurring beyond the acute phase of infection. Long COVID case status was determined solely from observed survey data and imputed values were not used for case ascertainment.

### Cohort Definition and Eligibility criteria

The cohort comprised participants who met the following criteria: 1) had evidence of SARS-CoV-2 infection, based on a positive PCR, antigen or serology test between December 01, 2020, and December 31, 2022. This infection date was defined as the index infection date in this study, ensuring for a pre-infection symptom assessment and a minimum of 12 months of post-infection follow-up; 2) had at least one follow-up assessment in the year before and after the index infection; 3) had an exact infection date or the imputation interval of no more than 90 days; 4) were identified as long COVID cases according to the primary long COVID definition; 5) were not previously identified as having long COVID prior to the index infection.

### Centroid-Based Partition Clustering Analysis of Longitudinal Symptoms

We applied K-means clustering for longitudinal data analysis, utilizing a centroid-based partitioning method. This approach assigns groups by minimizing the distance between each participant’s symptom trajectory within each cluster while maximizing the distance between cluster centroids ^33^. The method proceeded in two steps, as follows:

1. We first conducted hierarchical cluster analysis to estimate the number of optimal clusters by the Ward linkage method using McClain, Dunn, and Silhouette criteria ^14^. The McClain’s index, Dunn index, and Silhouette score assessed clustering quality by optimizing two key concepts: cluster compactness (how close points within a cluster are to each other) and cluster separation (how distinct clusters are from one another) ^34^.
2. We applied the K-means for longitudinal data clustering algorithm using the optimal cluster number determined in the step described above ^35^. Based on the selected optimal number of clusters, we performed longitudinal K-means consensus clustering with dynamic time warping distance to identify symptom clusters by counting each symptom equally ^36,37^. In this algorithm, each observation is reassigned to the group with the closest mean and the operation is repeated until convergence.

### Statistical Analysis

K-means clustering was performed separately on each of the 30 imputed datasets to assign participants into clusters based on longitudinal symptoms. To consolidate potentially inconsistent cluster assignments across imputations, we implemented a hierarchical aggregation procedure (the details are provided in **Appendix 3**) ^38–40^. This approach overcomes the limitations posed by arbitrary and potentially non-aligned cluster labels across imputations by relying on patterns of co-assignment rather than direct label matching. After finalizing the cluster assignments, we visualized symptom trajectories using radar plots and presented the proportions and corresponding 95% confidence intervals, stratified by clusters, symptom and time points (pre-infection, 3-, 6-, 9-, and 12-month post-infection). For each cluster and time point, pooled symptom proportions were calculated by averaging estimates across the 30 imputations, with the corresponding variances combined using Rubin’s rules to integrate both within- and between-imputation variability ^41,42^.

After participants were classified into clusters, we characterized the long COVID symptom burden by first computing, for each imputed dataset, the total number of symptoms at each time point (calculated as the sum of 15 binary symptoms). For each participant, the median symptom count across the 30 imputations was then calculated. For each cluster and time point, the pooled symptom burden was defined as the median of these participant-level medians, and the variability across imputations was calculated by computing the sample standard deviation of the medians divided by the square root of the number of participants, yielding approximate 95% confidence intervals as the pooled median ± 1.96 times this standard error ^41,42^.

We presented the prevalence of demographic, socioeconomic and comorbidities characteristics, prior infection status at index date, post index date reinfection status, and vaccination status, stratified by cluster. To examine factors associated with cluster assignment, we used multivariable logistic regression models to estimate adjusted odds ratios (aORs) and 95% CI for two comparisons: highest verse lowest symptom burden and moderate versus lowest symptom burden, with each model adjusted only for age and gender.

To further explore symptom variations within the highest symptom burden cluster, we conducted hierarchical clustering to identify distinct symptom phenotypes. The heatmap represents the prevalence of each symptom reported for at least six months post-infection across the identified phenotypic subgroups. Color intensity indicates symptom prevalence, with darker shades representing higher prevalence. Details of sensitivity analyses are provided in **Appendix 4**. We used SAS version 9.4 (SAS Institute, Cary, NC, USA) for data cleaning and R version 2024.09.0+375 (R Foundation for Statistical Computing, Vienna, Austria) for data analyses.

## RESULTS

### Long COVID Cases Characteristics

A total of 511 participants out of 1,787 infected individuals who met the eligibility criteria were included in the analysis (**eFigure 1**). Among these 511 long COVID cases, 22% were aged ≥50, 55% were female, 60% were White Non-Hispanic, 45% did not have a college degree, and 35% reported an annual household income of 35,000 or less (**Table 1**). In terms of comorbidities, 21% had 2 or more underlying conditions. The prevalence of specific conditions included mental health disorders (54%), type 2 diabetes (8%), heart disease (5.3%), HIV (5.1%), lung disease (4.7%), cancer (4.7%), kidney disease (2.0%), and immunodeficiency (7.2%). Regarding prior COVID-19 history, 30% of long COVID cases had a SARS-CoV-2 infection prior to the index infection date, and 18% experienced a subsequent SARS-CoV-2 infection during the 3-12 months following the index infection. Vaccination status preceding the index infection was as follows: 24% were unvaccinated, 5% were vaccinated but had not completed the primary series, 22% had completed the primary series, and 49% had completed the primary series and received at least one additional dose.

**Table 1:** Median Number of Long COVID Symptoms and 95% Confidence Intervals by Symptom Burden Cluster and Timepoint, the CHASING COVID Cohort Study (N = 511)

| Cluster | Timepoint | Median | Error bar |
| --- | --- | --- | --- |
| Lowest Symptom Burden | Pre-infection | 0 | -0.16,0.16 |
|  | 3-month post infection | 1 | 0.79,1.21 |
|  | 6-month post infection | 1 | 0.79,1.21 |
|  | 9-month post infection | 1 | 0.77,1.23 |
|  | 12-month post infection | 1 | 0.75,1.25 |
| Moderate Symptom Burden | Pre-infection | 1 | 0.76,1.24 |
|  | 3-month post infection | 3 | 2.73,3.27 |
|  | 6-month post infection | 3 | 2.65,3.35 |
|  | 9-month post infection | 2 | 1.69,2.31 |
|  | 12-month post infection | 3 | 2.61,3.39 |
| Highest Symptom Burden | Pre-infection | 4 | 3.59,4.41 |
|  | 3-month post infection | 5 | 4.51,5.49 |
|  | 6-month post infection | 6 | 5.56,6.44 |
|  | 9-month post infection | 5 | 4.51,5.49 |
|  | 12-month post infection | 6.25 | 5.73,6.77 |

### Symptoms Cluster Assignment

As shown in **Figure 1 and eTable 1**, distinct symptom profiles were observed across clusters. Among the individuals in Cluster 2, all of whom were identified as having long COVID, fatigue had the highest prevalence at 12 months post-infection (57%, 95% CI: 44-70%). Several other symptoms--difficulty running, difficulty walking, difficulty sleeping, gastrointestinal issues, headache, irritability, post-exertional malaise, myalgia, and trouble concentrating--each had a prevalence 46-53%, and collectively represented the most commonly reported symptoms within this cluster. For fatigue, the 12 months prevalence remained elevated compared to the pre-infection level of 51% (95% CI: 43-60%), though it was lower than the peak prevalence of 74% (95% CI: 63-85%) at 3-month post-infection.

**Figure 1.**
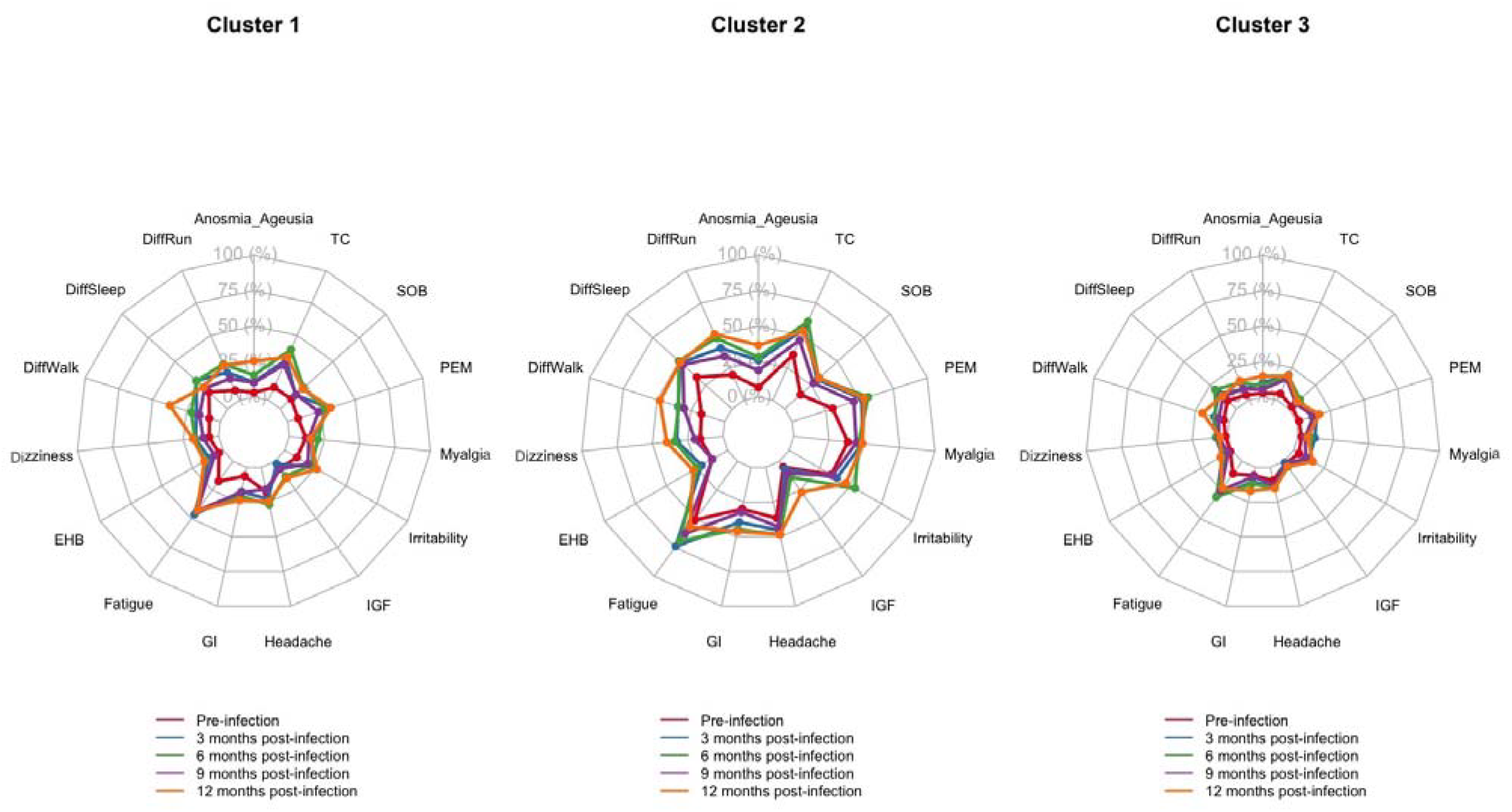
Radar plots representing the pooled prevalence estimates of Long COVID symptoms across three clusters at five time points, the CHASING COVID Cohort study, December 2020 – December 2023 (N=511) Abbreviations: PEM, post-exertional malaise; Anosmia/Ageusia, loss of smell or taste; DiffSleep, difficulty sleeping; LGF, low-grade fever; Irritabilt, irritability; DiffRun, difficulty running; DiffWalk, difficulty walking; SOB, shortness of breath; GI, gastrointestinal symptoms; EHB, erratic heartbeats; TC, trouble concentrating. Three side-by-side radar plots (Clusters 1–3) show the prevalence (0–100%) of 15 long-COVID symptoms—fatigue, trouble concentrating, post-exertional malaise, myalgia, difficulty sleeping, gastrointestinal symptoms, headache, irritability, dizziness, erratic heartbeats, shortness of breath, difficulty walking, difficulty running, loss/alteration of smell or taste, and low-grade fever—measured at five time points (pre-infection and 3, 6, 9, and 12 months post-infection). Cluster 2 has the highest and most persistent burden: multiple symptoms (especially trouble concentrating, fatigue, post-exertional malaise, sleep difficulty, GI symptoms, myalgia, and mobility limitations) reach roughly 40– 60% at 6–9 months and remain elevated at 12 months. Cluster 1 shows a moderate burden, with most symptoms around 20–35% and modest decline by 12 months. Cluster 3 shows the lowest burden, generally ∼10–25% across time. Pre-infection levels are lower than post-infection in all clusters.

In the Cluster 1, fatigue was the most prevalent symptom at 12-months post-infection (prevalence: 42%, 95% CI: 33-52%), with prevalence slightly declined compared to earlier post-infection time points (**Figure 1 and eTable 1**). In addition, post-exertional malaise, and trouble concentrating were prevalent close to 30% at 12-month post-infection, which remained elevated compared to their pre-infection level. In Cluster 3, although fatigue was the most prevalent at 12 months (24%, 95% CI: 16-31%), which was higher than pre-infection level and slightly declined compared to 3-, 6-, and 9-months post-infection levels. The prevalence of difficulty sleeping was slightly declined at month 12 compared to the 3-, 6-, and 9-months post-infection.

As shown in **Table 1**, the median number of long COVID symptoms across time points was used to characterize the Cluster 2, 1, 3 as highest, moderate, and lowest symptom burden clusters, respectively. The highest symptom burden cluster exhibited a consistently elevated symptom profile, with median symptom counts of 5–6 throughout the post-infection period. The cluster featured by moderate symptom burden had a median of 3 symptoms at 3-, 6- and 12-months post-infection, with a slight decline to 2 symptoms at 9 months. In contrast, the lowest symptom burden cluster maintained a stable median of 1 symptom across all post-infection time points.

### Risk Factors Associated with Cluster Assignment

Adjusted odds ratios (aOR) for classification into highest vs. lowest symptom burden clusters were shown in **Table 2**. Participants aged over 50 had more than a two-fold adjusted odds of being classified into the highest (vs. lowest) symptom burden cluster compared to those under 50 (aOR 2.68, 95% CI: 1.59-4.53). Female participants had significantly higher adjusted odds of being classified into the highest (vs. lowest) symptom burden cluster compared to males (aOR 2.36, 95% CI: 1.48-3.76). Additionally, individuals with two or more pre-infection comorbidities had over three times the adjusted odds of being classified into the highest (vs. lowest) symptom burden cluster compared to those with no comorbidities (aOR 5.91, 95% CI: 3.08-11.37). Participants with a history of mental health disorders had significantly elevated odds of being classified into the highest (vs. lowest) symptom burden cluster (aOR: 2.65, 95% CI: 1.65–4.25), and those with immunodeficiency had more than four-fold increased odds (aOR: 4.23, 95% CI: 1.71–10.51).

**Table 2:** Characteristics and Adjusted Odds of Classification into Long COVID Symptom Burden Clusters Identified by K-Means Clustering, the CHASING COVID Cohort Study, December 2020 – December 2023.

|  | Total | Lowest symptom burden | Moderate symptom burden | Highest symptom burden | Moderate vs. Lowest Symptom Burden Cluster | Highest vs. Lowest Symptom Burden Cluster |
| --- | --- | --- | --- | --- | --- | --- |
|  | N=511 | N = 199 | N = 174 | N = 138 | aOR (95% CI) | aOR (95% CI) |
| **Age** |  |  |  |  |  |  |
| ≤50 | 398 (78%) | 166 (83%) | 141 (81%) | 91 (66%) | ref | ref |
| >50 | 113 (22%) | 33 (17%) | 33 (19%) | 47 (34%) | 1.23 (0.72 - 2.1) | **2.68 (1.59 - 4.53)** |
| **Gender** |  |  |  |  |  |  |
| Male | 210 (41%) | 100 (50%) | 68 (39%) | 42 (30%) | ref | ref |
| Female | 282 (55%) | 95 (48%) | 95 (55%) | 92 (67%) | 1.48 (0.97 - 2.25) | **2.36 (1.48 - 3.76)** |
| Non-binary | 19 (3.7%) | 4 (2.0%) | 11 (6.3%) | 4 (2.9%) | 4.14 (0.86 - 13.55) | 2.74 (0.65 - 11.62) |
| **Race/Ethnicity** |  |  |  |  |  |  |
| White Non-Hispanic | 307 (60%) | 115 (58%) | 102 (59%) | 90 (65%) | ref | ref |
| Hispanic | 92 (18%) | 44 (22%) | 31 (18%) | 17 (12%) | 0.83 (0.48 - 1.43) | 0.62 (0.32 - 1.2) |
| Black Non-Hispanic | 61 (12%) | 24 (12%) | 25 (14%) | 12 (8.7%) | 1.24 (0.66 - 2.32) | 0.7 (0.32 - 1.51) |
| Asian Non-Hispanic | 28 (5.5%) | 12 (6.0%) | 7 (4.0%) | 9 (6.5%) | 0.63 (0.23 - 1.7) | 1.21 (0.47 - 3.09) |
| Other Non-Hispanic | 23 (4.5%) | 4 (2.0%) | 9 (5.2%) | 10 (7.2%) | 2.54 (0.75 - 8.6) | 3.83 (0.93 - 13) |
|  | N=511 | N = 199 | N = 174 | N = 138 | aOR (95% CI) | aOR (95% CI) |
| **Household income** |  |  |  |  |  |  |
| ≤35k or unknown | 180 (35%) | 61 (31%) | 66 (38%) | 53 (38%) | 1.29 (0.74 - 2.26) | 1.31 (0.71 - 2.41) |
| 35-100k | 217 (42%) | 90 (45%) | 71 (41%) | 56 (41%) | 0.98 (0.57 - 1.67) | 0.96 (0.53 - 1.72) |
| ≥100k | 114 (22%) | 48 (24%) | 37 (21%) | 29 (21%) | ref | ref |
| **Education** |  |  |  |  |  |  |
| Less than college | 229 (45%) | 75 (38%) | 84 (48%) | 70 (51%) | ref | ref |
| College and above | 282 (55%) | 124 (62%) | 90 (52%) | 68 (49%) | 0.68 (0.45 - 1.03) | 0.64 (0.41 - 1.01) |
| **SARS-CoV-2 infections prior to index SARS-CoV-2** |  |  |  |  |  |  |
| No | 357 (70%) | 141 (71%) | 118 (68%) | 98 (71%) | ref | ref |
| Yes | 154 (30%) | 58 (29%) | 56 (32%) | 40 (29%) | 1.27 (0.81 - 1.99) | 1.16 (0.7 - 1.9) |
| **Post index infection date SARS-CoV-2 infection** |  |  |  |  |  |  |
| No | 420 (82%) | 166 (83%) | 143 (82%) | 111 (80%) | ref | ref |
| Yes | 91 (18%) | 33 (17%) | 31 (18%) | 27 (20%) | 1.15 (0.67 - 1.97) | 1.32 (0.74 - 2.36) |
| **Any COVID-19 vaccination in the past 6 months** |  |  |  |  |  |  |
| No | 216 (42%) | 87 (44%) | 74 (43%) | 55 (40%) | ref | ref |
| Yes | 295 (58%) | 112 (56%) | 100 (57%) | 83 (60%) | 1.09 (0.71 - 1.65) | 1.32 (0.83 - 2.09) |
| **Vaccination status before infection** |  |  |  |  |  |  |
| No Vaccine | 121 (24%) | 47 (24%) | 42 (24%) | 32 (23%) | ref | ref |
| Vaccinated but not completing the primary series | 28 (5.5%) | 10 (5.0%) | 11 (6.3%) | 7 (5.1%) | 1.3 (0.5 - 3.4) | 1.11 (0.37 - 3.35) |
| Completed the primary series | 111 (22%) | 48 (24%) | 32 (18%) | 31 (22%) | 0.77 (0.42 - 1.43) | 0.99 (0.51 - 1.91) |
| Completed the primary series and at least one additional dose | 251 (49%) | 94 (47%) | 89 (51%) | 68 (49%) | 1.1 (0.65 - 1.84) | 1.16 (0.66 - 2.05) |
| **Smoking cigarettes or e-cigarettes** |  |  |  |  |  |  |
| No | 336 (66%) | 138 (69%) | 114 (66%) | 84 (61%) | ref | ref |
| Yes | 175 (34%) | 61 (31%) | 60 (34%) | 54 (39%) | 1.23 (0.79 - 1.92) | 1.66 (0.93 - 2.66) |
| **Comorbidity of Obesity** |  |  |  |  |  |  |
| No | 335 (66%) | 138 (69%) | 107 (61%) | 90 (65%) | ref | ref |
| Yes | 176 (34%) | 61 (31%) | 67 (39%) | 48 (35%) | 1.34 (0.87 - 2.08) | 1.01 (0.63 - 1.64) |
| **Comorbidity of Cancer** |  |  |  |  |  |  |

|  | **Total** | **Lowest symptom burden** | **Moderate symptom burden** | **Highest symptom burden** | **Moderate vs. Lowest Symptom Burden Cluster** | **Highest vs. Lowest Symptom Burden Cluster** |
| --- | --- | --- | --- | --- | --- | --- |
|  | **N=511** | **N = 199** | **N = 174** | **N = 138** | **aOR (95% CI)** | **aOR (95% CI)** |
| No | 487 (95%) | 191 (96%) | 169 (97%) | 127 (92%) | ref | ref |
| Yes | 24 (4.7%) | 8 (4.0%) | 5 (2.9%) | 11 (8.0%) | 0.68 (0.21 - 2.15) | 1.64 (0.61 - 4.42) |
| **Comorbidity of Kidney Disease** |  |  |  |  |  |  |
| No | 501 (98%) | 198 (99%) | 173 (99%) | 130 (94%) | ref | ref |
| Yes | 10 (2.0%) | 1 (0.5%) | 1 (0.6%) | 8 (5.8%) | 0.94 (0.06 - 15.78) | 5.6 (0.65 - 47.92) |
| **Comorbidity of Lung Disease** |  |  |  |  |  |  |
| No | 487 (95%) | 193 (97%) | 167 (96%) | 127 (92%) | ref | ref |
| Yes | 24 (4.7%) | 6 (3.0%) | 7 (4.0%) | 11 (8.0%) | 1.18 (0.38 - 3.64) | 2.1 (0.73 - 6.05) |
| **Comorbidity of Type-2 Diabetes** |  |  |  |  |  |  |
| No | 470 (92%) | 184 (92%) | 166 (95%) | 120 (87%) | ref | ref |
| Yes | 41 (8.0%) | 15 (7.5%) | 8 (4.6%) | 18 (13%) | 0.64 (0.26 - 1.57) | 1.95 (0.9 - 4.23) |
| **Comorbidity of Heart Disease** |  |  |  |  |  |  |
| No | 484 (95%) | 189 (95%) | 166 (95%) | 129 (93%) | ref | ref |
| Yes | 27 (5.3%) | 10 (5.0%) | 8 (4.6%) | 9 (6.5%) | 1.02 (0.39 - 2.69) | 1.4 (0.53 - 3.71) |
| **Comorbidity of HIV** |  |  |  |  |  |  |
| No | 485 (95%) | 188 (94%) | 164 (94%) | 133 (96%) | ref | ref |
| Yes | 26 (5.1%) | 11 (5.5%) | 10 (5.7%) | 5 (3.6%) | 1.25 (0.5 - 3.12) | 0.93 (0.3 - 2.87) |
| **Comorbidity of Mental health disorders** |  |  |  |  |  |  |
| No | 237 (46%) | 112 (56%) | 80 (46%) | 45 (33%) | ref | ref |
| Yes | 274 (54%) | 87 (44%) | 94 (54%) | 93 (67%) | 1.38 (0.91 - 2.11) | **2.65 (1.65 - 4.25)** |
| **Comorbidity of Immunodeficiency** |  |  |  |  |  |  |
| No | 474 (93%) | 192 (96%) | 165 (95%) | 117 (85%) | ref | ref |
| Yes | 37 (7.2%) | 7 (3.5%) | 9 (5.2%) | 21 (15%) | 1.4 (0.5 - 3.87) | **4.23 (1.71 - 10.51)** |
| **Number of comorbidities** |  |  |  |  |  |  |
| 0 | 184 (36%) | 87 (44%) | 66 (38%) | 31 (22%) | ref | ref |
| 1 | 222 (43%) | 88 (44%) | 81 (47%) | 53 (38%) | 1.09 (0.69 - 1.7) | 1.52 (0.88 - 2.63) |
| >=2 | 105 (21%) | 24 (12%) | 27 (16%) | 54 (39%) | 1.4 (0.74 - 2.67) | **5.91 (3.08 - 11.37)** |

### Symptom Phenotype for those Classified in the Highest Symptom Burden Cluster

Figure 2. illustrates the distribution of symptom phenotypes among participants in the highest symptom burden cluster, with symptoms categorized into three phenotypes (A, B, and C), each representing approximately one-third of the cluster (32%, 36%, and 32%, respectively). The heatmap displays the prevalence of specific symptoms that were reported at multiple time points spanning six or more months post-infection, with darker shades indicating higher prevalence. Phenotype A is predominantly characterized by multi-systemic dysfunction, with a higher prevalence of fatigue, post-exertional malaise, and brain fog/trouble concentrating. Phenotype B is defined by a combination of psychiatric and neurological symptoms, including irritability and difficulty sleeping. Phenotype C is primarily associated with physical and respiratory limitations, including fatigue, difficulty walking and running, and shortness of breath.

**Figure 2.**
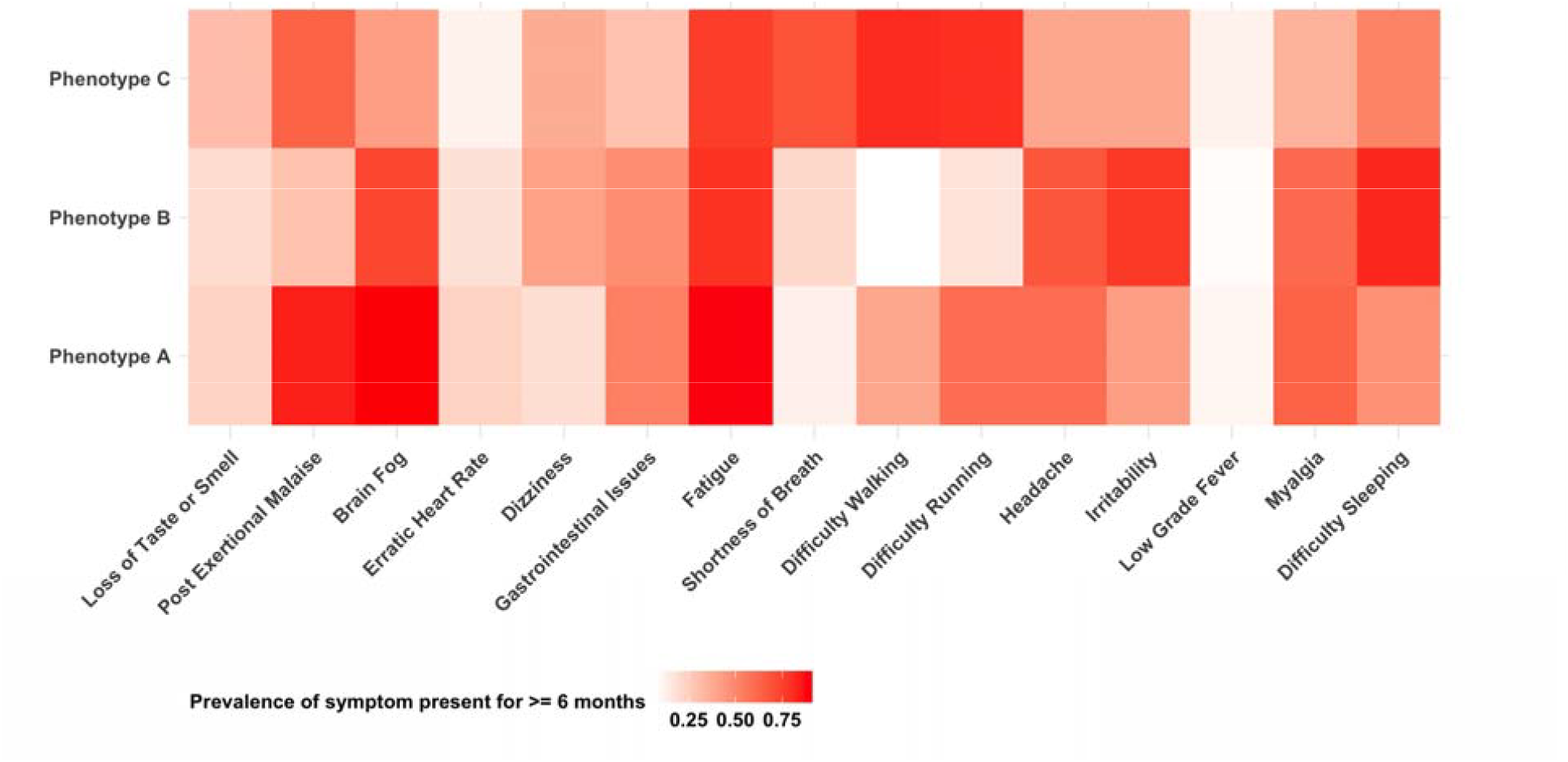
Symptom Phenotypes Among Individuals with Highest Symptom Burden Figure Description: Symptom phenotypes among individuals classified in the high symptom burden cluster, showing the prevalence of symptoms persisting for ≥6 months. •Phenotype A: Predominantly neurological and multi-systematic dysfunction, including a higher prevalence of fatigue, post-exertional malaise, and brain fog/trouble concentrating. •Phenotype B: Defined by a mix of psychiatric and neurological symptoms, including irritability and difficulty sleeping. •Phenotype C: Characterized by physical and respiratory limitations, including fatigue, difficulty walking and running, and shortness of breath.

## DISCUSSION

We identified distinct long COVID symptom burden clusters up to 12 months post-infection in a community-based cohort, demonstrating heterogeneity in symptom counts, profile-specific recovery trajectories, and associated risk factors. Unlike EHR/clinic-based studies, systematic, repeated community self-reporting captured individuals who did not seek care, improving case ascertainment and generalizability. These findings clarify the spectrum of long COVID presentations and can inform diagnosis, risk stratification, and targeted interventions.

Using a data-driven approach, we delineated three long COVID subgroups—highest, moderate, and lowest symptom burden—defined by both total symptom counts and longitudinal prevalence profiles, capturing differences in overall intensity across time points. This structure accords with prior reports of heterogeneity ^43^ and, consistent with RECOVER ^7^, includes a subset with persistently elevated burden. In the high-burden cluster, fatigue, concentration difficulties, post-exertional malaise, and gastrointestinal symptoms predominated, with prevalence peaking at ∼6 months. Across the 12-month follow-up we observed a descriptive decline in fatigue in all clusters, which may indicate partial improvement; however, because clusters were formed on elevated early symptoms, some of this trend could reflect regression to the mean rather than true biological recovery and should be interpreted cautiously. Overall, these findings reinforce the diversity of long-COVID symptomatology and trajectories and support tailored management, risk stratification, and targeted intervention development ^44^.

Consistent with prior reports, we found that older age, female sex, and the presence of underlying conditions such as mental health disorders and immunodeficiency were associated with an increased risk of being in the highest versus the lowest symptom burden cluster ^45,46^. Recent studies have identified factors associated with varying symptom burdens among individuals with long COVID. For instance, Evans et al. indicated that female sex and older age were linked to more severe long COVID symptom profiles ^46^. In a large cohort study, Xie et al. found that individuals with baseline mental health conditions, such as anxiety and depression, had a significantly higher risk of experiencing persistent long COVID symptoms ^47^. Similarly, Al-Aly et al. demonstrated that individuals with compromised immune function were at increased risk for prolonged and severe symptomatology following acute SARS-CoV-2 infection, likely due to impaired viral clearance and ongoing immune activation ^48^. These studies support the hypothesis that underlying psychosocial and immunological vulnerability may contribute to more severe or persistent long COVID phenotypes. These findings suggest that targeted interventions may be necessary for these subgroups to address their specific needs. To be noted, this study incorporated key social determinants of health—including education and household income—which are often underrepresented in long COVID research. Although these variables were not significantly associated with symptom burden in our primary models, their inclusion provides important context and may help explain disparities in long COVID outcomes in future studies.

In the highest symptom burden cluster, the identification of distinct phenotypes—including multi-systematic symptoms, neurological and psychiatric symptoms, and physical and respiratory limitations—underscored the heterogeneity of long COVID manifestations. This variability had been corroborated by multiple studies, emphasizing the necessity for personalized care strategies. A multicenter, prospective cohort study identified four distinct phenotypes of post-COVID conditions at 3 and 6 months, including minimal symptoms, tiredness/headache/musculoskeletal symptoms, loss of smell/taste, and multisystem symptoms ^49^. Blankestijn et al. delineated three distinct long COVID clusters based on patient characteristics, lung function, and symptom severity, further illustrating the complexity of the condition ^50^. These findings collectively suggested that the observed heterogeneity in long COVID symptomatology might reflect different mechanisms and pathophysiologic pathways. Understanding these distinct symptom profiles could potentially inform more precise diagnostic criteria, improve risk stratification, and ultimately guide the development of more effective, phenotype-specific therapeutic strategies.

This study had several important limitations. Symptoms were self-reported rather than clinician-elicited, which may overestimate prevalence; however, uniform ascertainment across participants and time points limits differential bias and preserves comparative validity. Symptom burden was quantified by counts rather than severity, constraining inferences about intensity; future work should capture severity. Follow-up requirements may have excluded those with the most debilitating illness, potentially underrepresenting the highest-burden cluster. Finally, post–time-zero SARS-CoV-2 reinfections could have contributed symptoms not attributable to the index infection, though reinfection rates were balanced across clusters, making differential effects unlikely.

This study highlighted the heterogeneity of long COVID symptom trajectories and its associated risk factors. The identification of distinct symptom sub-phenotypes underscored the complexity of long COVID and the need for personalized approaches to management and care. Recognizing the diverse trajectories of recovery and symptom patterns is essential for diagnosis, improving outcomes and addressing the long-term impact of this condition.

## Data Availability

The data underlying this study contain potentially identifiable health information and are not publicly available due to ethical and legal restrictions. De-identified data may be shared on reasonable request and subject to institutional approvals and a Data Use Agreement. Requests will be reviewed case-by-case by the study investigators and the CUNY ISPH data governance/IRB. Please contact the corresponding author.

https://cunyisph.org/chasing-covid/

## FUNDING

Funding for this project is provided by The National Institute of Allergy and Infectious Diseases (NIAID), award number UH3AI133675 (MPIs: D Nash and C Grov), Pfizer Inc, the CUNY Institute for Implementation Science in Population Health (cunyisph.org) and the COVID-19 Grant Program of the CUNY Graduate School of Public Health and Health Policy.

The NIH played no role in the production of this manuscript nor necessarily endorses the findings. The study design was developed by CUNY without input from Pfizer.

## PATIENT CONSENT STATEMENT

Informed consent forms were completed in a web browser on participants’ computer or mobile device at baseline, each round of serological testing, and at periodic follow-up assessments. The study was approved by the Institutional Review Boards of the City University Of New York (CUNY) Graduate School of Public Health and Health Policy (New York, NY, U.S.) (protocol 2020-0256).

