## Supplementary files for "Long COVID Longitudinal Symptoms Burden Clusters Within A National Community-Based Cohort"

**Supplementary File**

### Appendix Table 1: STROBE (Strengthening the Reporting of Observational studies with Epidemiology) Checklist of items that should be included in reports of cohort studies.

|  | Item | Recommendation | Page |
| --- | --- | --- | --- |
| **Title and abstract** | 1 | (a) Indicate the study’s design with a commonly used term in the title or the abstract |  |
|  |  | (b) Provide in the abstract an informative and balanced summary of what was done and what was found | 1-2 |
| Introduction | | | |
| Background/rationale | 2 | Explain the scientific background and rationale for the investigation being reported | 3 |
| Objectives | 3 | State specific objectives, including any prespecified hypotheses | 3 |
| Methods | | | |
| Study design | 4 | Present key elements of study design early in the paper | 3-4 |
| Setting | 5 | Describe the setting, locations, and relevant dates, including periods of recruitment, exposure, follow-up, and data collection | 3-4 |
| Participants | 6 | (a) Give the eligibility criteria, and the sources and methods of selection of participants. Describe methods of follow-up |  |
|  |  | (b) For matched studies, give matching criteria and number of exposed and unexposed | 5 |
| Variables | 7 | Clearly define all outcomes, exposures, predictors, potential confounders, and effect modifiers. Give diagnostic criteria, if applicable | 4, 5 |
| Data sources/ measurement | 8 | For each variable of interest, give sources of data and details of methods of assessment (measurement). Describe comparability of assessment methods if there is more than one group | 3, 4, 5 |
| Bias | 9 | Describe any efforts to address potential sources of bias | 4, Appendix 1 & 4 |
| Study size | 10 | Explain how the study size was arrived at | eFigure 1 |
| Quantitative variables | 11 | Explain how quantitative variables were handled in the analyses. If applicable, describe which groupings were chosen and why | 4, 5, Appendix 1, 2 |
| Statistical methods | 12 | (a) Describe all statistical methods, including those used to control for confounding |  |
|  |  | (b) Describe any methods used to examine subgroups and interactions | 5, 6, Appendix 3, 4, eTable 2 |
|  |  | (c) Explain how missing data were addressed |  |
|  |  | (d) If applicable, explain how loss to follow-up was addressed |  |
|  |  | (e) Describe any sensitivity analyses |  |
| Results | | |  |
| Participants | 13 | (a) Report numbers of individuals at each stage of study—eg numbers potentially eligible, examined for eligibility, confirmed eligible, included in the study, completing follow-up, and analyzed | 6, eFigure 1 |
|  |  | (b) Give reasons for non-participation at each stage |  |
|  |  | (c) Consider use of a flow diagram |  |
| Descriptive data | 14 | (a) Give characteristics of study participants (eg demographic, clinical, social) and information on exposures and potential confounders | 6-7, Table 2 |
|  |  | (b) Indicate number of participants with missing data for each variable of interest |  |
|  |  | (c) Summarize follow-up time (eg, average and total amount) |  |
| Outcome data | 15 | Report numbers of outcome events or summary measures over time | 7, Figure 1, Table 1, eTable 1 |
| Main results | 16 | (a) Give unadjusted estimates and, if applicable, confounder-adjusted estimates and their precision (eg, 95% confidence interval). Make clear which confounders were adjusted for and why they were included  (b) Report category boundaries when continuous variables were categorized  (c) If relevant, consider translating estimates of relative risk into absolute risk for a meaningful time period | 7, Table 2 |
| Other analyses | 17 | Report other analyses done—eg analyses of subgroups and interactions, and sensitivity analyses | 8, Figure 2, eFigure 2, eTable 3 |
| **Discussion** |  |  |  |
| Key results | 18 | Summarise key results with reference to study objectives | 8 |
| Limitations | 19 | Discuss limitations of the study, taking into account sources of potential bias or imprecision. Discuss both direction and magnitude of any potential bias | 9 |
| Interpretation | 20 | Give a cautious overall interpretation of results considering objectives, limitations, multiplicity of analyses, results from similar studies, and other relevant evidence | 8-9 |
| Generalisability | 21 | Discuss the generalisability (external validity) of the study results | 9 |
| **Other information** |  |  |  |
| Funding | 22 | Give the source of funding and the role of the funders for the present study and, if applicable, for the original study on which the present article is based | 10 |

### Appendix 1: Determination of SARS-CoV-2 Infection Status and Dates

The infection status was determined by: (1) self-reported testing positive on a Polymerase Chain Reaction (PCR) test or a rapid antigen test, whether conducted by a healthcare provider or using a home-based rapid test; or (2) testing positive for total antibodies (IgA, IgM, or IgG) to the SARS-CoV-2 nucleocapsid protein using dried blood spot (DBS) samples collected in the study; or (3) meeting the Council of State and Territorial Epidemiologists (CSTE) criteria for probable case definition, which was applied only during December 6, 2021 to January 11, 2022 when the U.S. testing capacity was limited during the initial Omicron surge ^1–4^. Participants were considered probable cases if they reported: (1) either one key symptom (cough, shortness of breath, altered smell/taste, or chest pain) or at least two other symptoms (fever, chills, myalgia, headache, sore throat, nausea/vomiting, or congestion); and (2) epidemiologic linkage within 14 days, such as close contact with a case or working in healthcare ^5^.

*Determination of SARS-CoV-2 Infection Dates*

Infection dates occurred within a known interval but the exact time within the interval was unknown. Exact infection dates were imputed within the known interval, and the imputation was based on how the infection was identified: self-reported positive PCR or antigen tests, serology-confirmed infections, or probable case per the CSTE criteria.

(1) Infection Identified by Positive PCR or Antigen Tests: At each follow-up, participants were prompted to provide exact dates of their positive PCR or antigen tests, and when possible, we used the exact date reported If participants provided only the month, the midpoint of that month was assigned as the infection date. For participants who did not report a month of infection, the infection date was imputed as the midpoint of the interval between the survey where they reported the infection and the previous follow-up assessment where they had no evidence of SARS-CoV-2 infection. Only infection with exact dates or those imputed using the midpoint approach with an interval of less than 90 days were included. Among infections identified by self-reported positive PCR or antigen tests, 72% were confirmed by a study-collected DBS sample within one year of infection.

(2) Infections Identified by Positive Serology Tests: Participants were invited to complete serologic testing using an at-home self-collected dried blood spot (DBS) specimen collection kit during three periods—from April through September 2020 (Serology Period 1), November 2020 through March 2021 (Serology Period 2), and March 2022 through June 2022 (Serology Period 3) ^6^. For serology-identified infections accompanied by a self-reported positive PCR or antigen test, or meeting CSTE criteria, infection dates were determined using the imputation intervals described in the relevant sections. Serology-identified infections without a prior viral or serology test or CSTE evidence were excluded, as infection dates could not be reliably estimated within a 90-day interval.

(3) Infections Identified by CSTE Criteria: For infections identified based on epidemiologic linkage and symptoms consistent with COVID-19, infection dates were assigned as the midpoint of the interval between the assessment date and 14 days prior, as the epidemiologic linkage was assessed within two weeks prior to the follow-up assessment. Among these cases, 57% were confirmed by a study-collected DBS sample within one year.

### Appendix 2: Demographics, Socioeconomics, Comorbidities, Prior Infections, and COVID-19 Vaccination Status

Demographic data of age, gender, race/ethnicity, education, household income, obesity status, smoking history and number of comorbidities were collected at enrollment between March and July 2020 and were available for all participants. Obesity was defined according to CDC criteria as a Body Mass Index (BMI) of ≥30.0 ^7^. Participants who reported smoking cigarettes or e-cigarette products every day or some days were classified as smokers. The number of comorbidities was counted from ten self-reported physician-diagnosed chronic systemic health conditions, including current asthma, cancer, kidney diseases, lung diseases, liver diseases, type-2 diabetes, heart conditions, HIV/AIDS, mental health disorders, and immunosuppression.

The employment category (employed, out of work, retired, home maker, students), geographic region of residence (northeast, Midwest, south, west), and material hardship were collected at each follow-up assessment approximately every three months. Participants experiencing either food insecurity or housing instability were classified as having material hardship. Food insecurity was assessed using the United States Department of Agriculture (USDA) Household Food Security Survey (HFSS) ^8^. Participants responded, “often true”, “sometimes true”, or “never true” to three statements: “We couldn’t afford to eat balanced meals”, “We worried whether our food would run out before we got money to buy more”, and “The food that we bought just didn’t last, and we didn’t have money to get more”. Those answering “often true” or “sometimes true” to any of these three statements were classified as food insecure ^9^. Housing instability was assessed with a question adapted from the Behavioral Risk Factor Surveillance System survey (BRFSS), asking how often participants worried about affording rent or mortgage payments ^10^. Those who answered “Always” or “Usually” were considered housing unstable ^11^.

In addition, because participants could experience repeated SARS-CoV-2 infections, we defined the prior SARS-CoV-2 at the time of the index infection as any documented infection occurring at least 90 days before the index date ^12^. Reinfection after the index date was defined as any subsequent infection occurring between 3 and 12 months following the index infection. COVID-19 vaccination history was determined based on self-reported vaccination records that were collected at each follow up assessment since December 2020 ^13^. Any dose of a COVID-19 vaccine administered within 6 months prior to the index infection date was classified as pre-infection vaccination, regardless of vaccine type or manufacturer.

### Appendix 3: The steps of final cluster assignment

First, the cluster assignment results from all imputations were merged into a single wide-format matrix, where each row represented a participant and each column denoted the cluster assignment from one of the 30 imputations. Second, we computed a co-occurrence matrix by calculating, for each pair of participants, the proportion of imputations in which they were assigned to the same cluster. Third, the co-occurrence matrix was transformed into a distance matrix by subtracting each value from one, such that smaller values represented more consistent co-clustering. Fourth, hierarchical clustering using average linkage was applied to this distance matrix to construct a dendrogram, where participants were successively grouped based on their average pairwise distances. Fifth, the dendrogram was cut at a level yielding the desired number of final clusters—in this study, three—thereby assigning each participant a single, consolidated cluster label that reflected their co-clustering patterns across imputations.

### Appendix 4: Sensitivity analysis plan

To explore variations in long COVID case definitions and assess the robustness of our findings, we conducted sensitivity analyses using two alternative definitions. The first alternative definition required at least one symptom to be present at two distinct time points: an initial occurrence between 3 and 10 months post-infection, followed by a second occurrence at least 60 days later, but no later than 12 months post-infection. The symptoms at these two time points were not required to be the same, but both had to be absent in the year preceding the index infection date. The second alternative definition classified cases based on self-identified long COVID, defined as “experiencing symptoms more than 4 weeks after you first had COVID-19 that are not explained by something else?". This question was from the U.K.’s Office for National Statistics (ONS) COVID-19 infection survey and was collected in each follow-up assessment since June 2022 ^14,15^.

We conducted the following sensitivity analysis: (1) We used a Finite Mixture Model of Latent Class Analysis (LCA) to derive the clusters as an alternative model-based approach to clustering, which provided a probabilistic framework for identifying latent subgroups in the data ^16–18^. (2) To evaluate the sensitivity of clusters assignments to variation in long COVID case definitions, we re-defined the analytical cohort based on the two alternative long COVID case definitions previously described. (3) To evaluate the robustness of symptom clustering results, we conducted a sensitivity analysis by excluding individuals who had evidence of any prior SARS-CoV-2 infection before their index SARS-CoV-2 infection date. This exclusion criterion was implemented to ensure that the post-infection symptom trajectories analyzed were attributable solely to the index infection, without influence from earlier infections that may have contributed to cumulative or residual symptom burden. Prior infections could confound symptom presentation and progression, making it more difficult to attribute post-acute symptoms specifically to the index infection. By restricting the sample to individuals with no documented history of SARS-CoV-2 infection before the index event, we aimed to reduce misclassification and better isolate the effect of a single infection episode on long-term symptom patterns.

### eFigure 1: Flow chart of participants' inclusion and exclusion process for the analytical sample

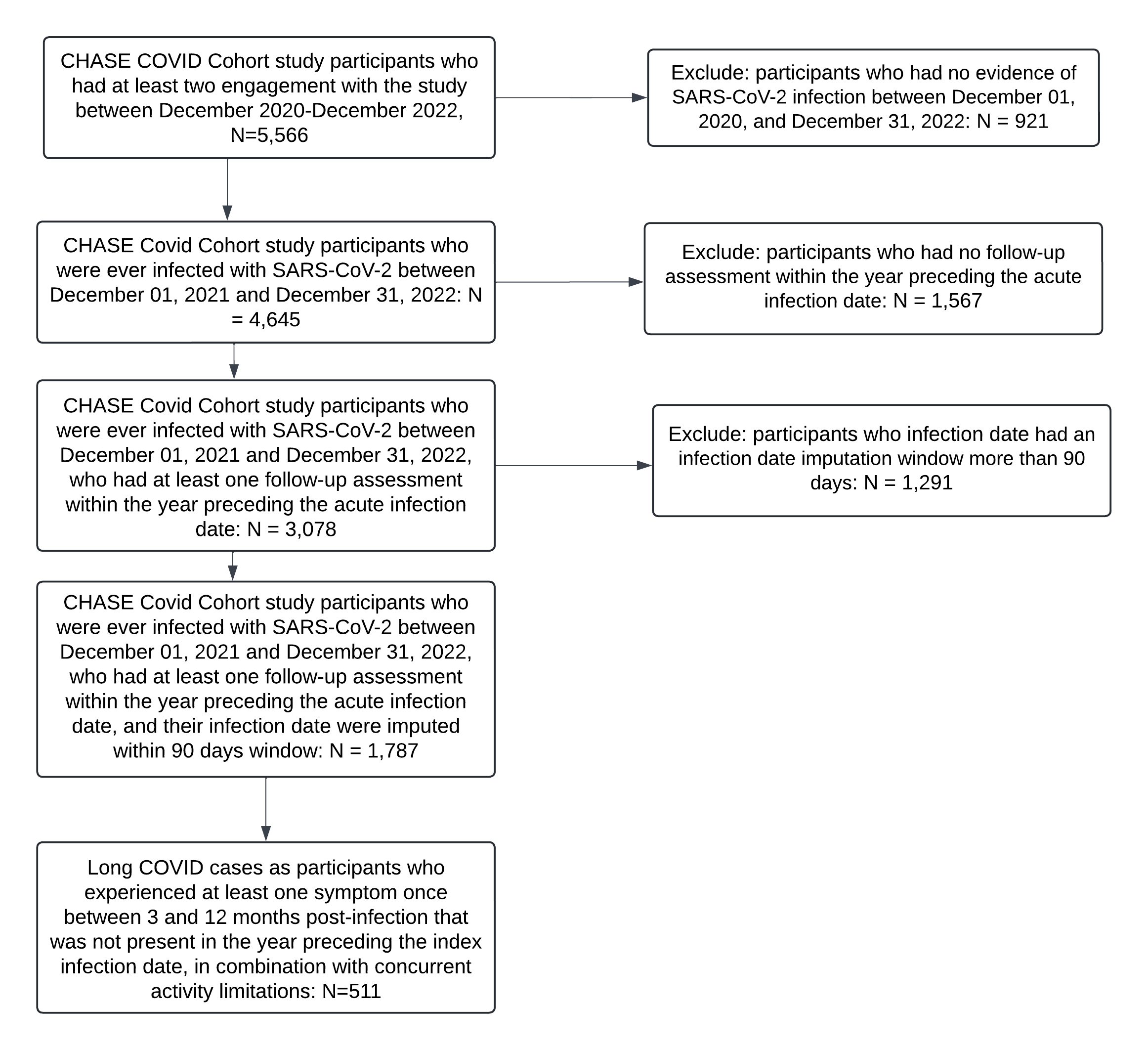

### eFigure 2: For each sensitivity analysis, the radar plots representing the prevalence of Long COVID symptoms across three clusters at five different time points, the CHASING COVID Cohort study, December 2020 – December 2023

a. **Sensitivity analysis using Latent Class Analysis (LCA)**

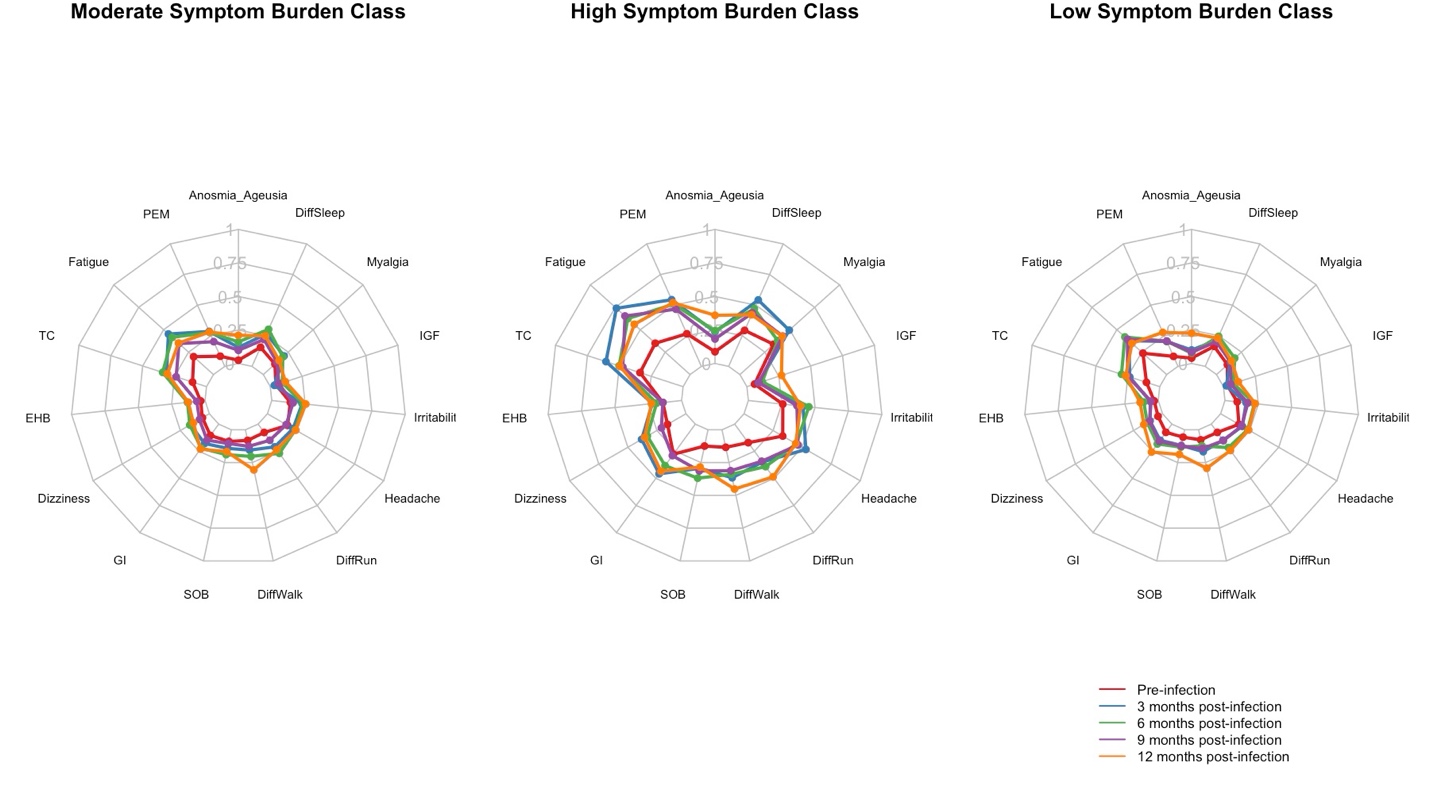

b. **Sensitivity analysis defining long COVID cases as at least one symptom to be present at two distinct time points**

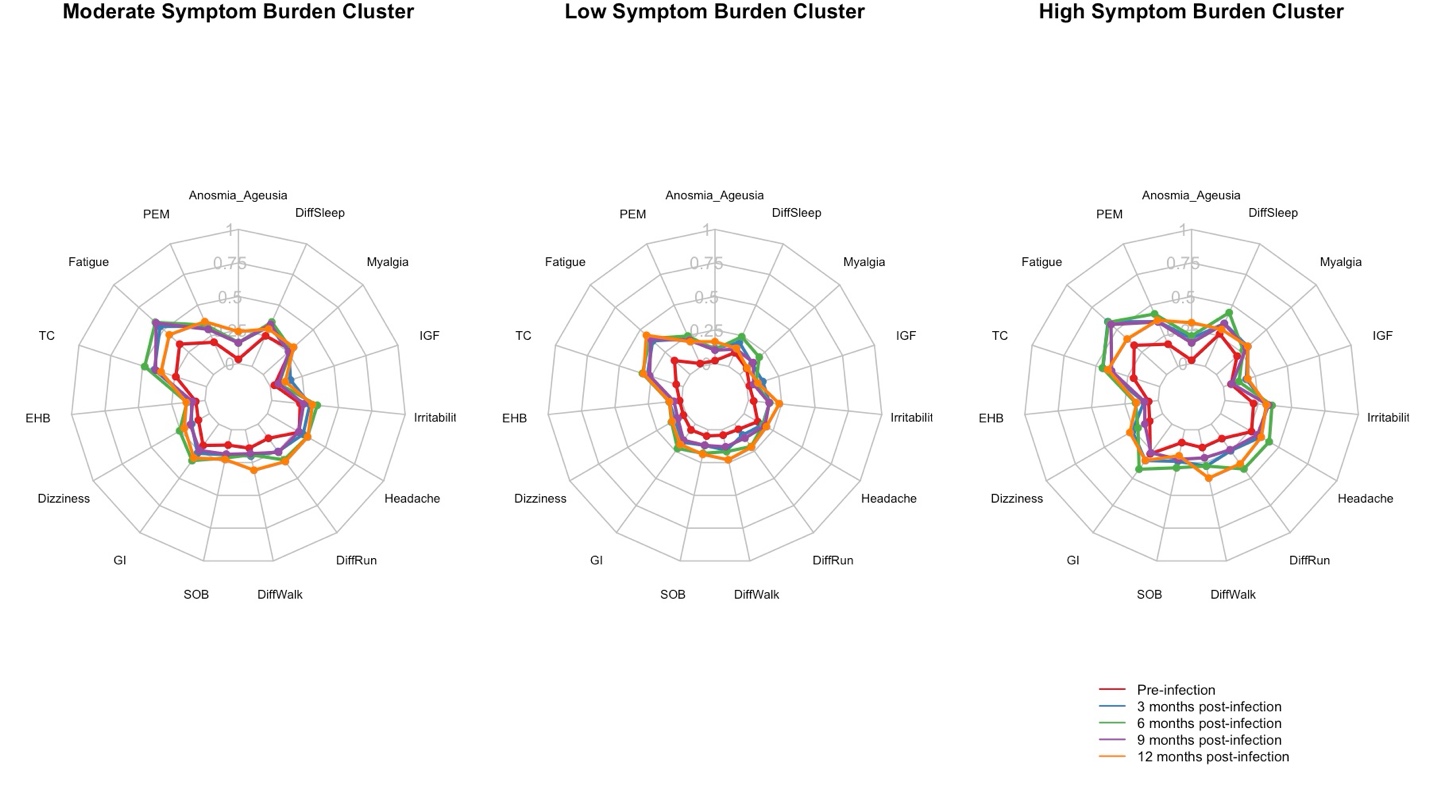

c. **Sensitivity analysis defining long COVID cases based on self-identified long COVID**

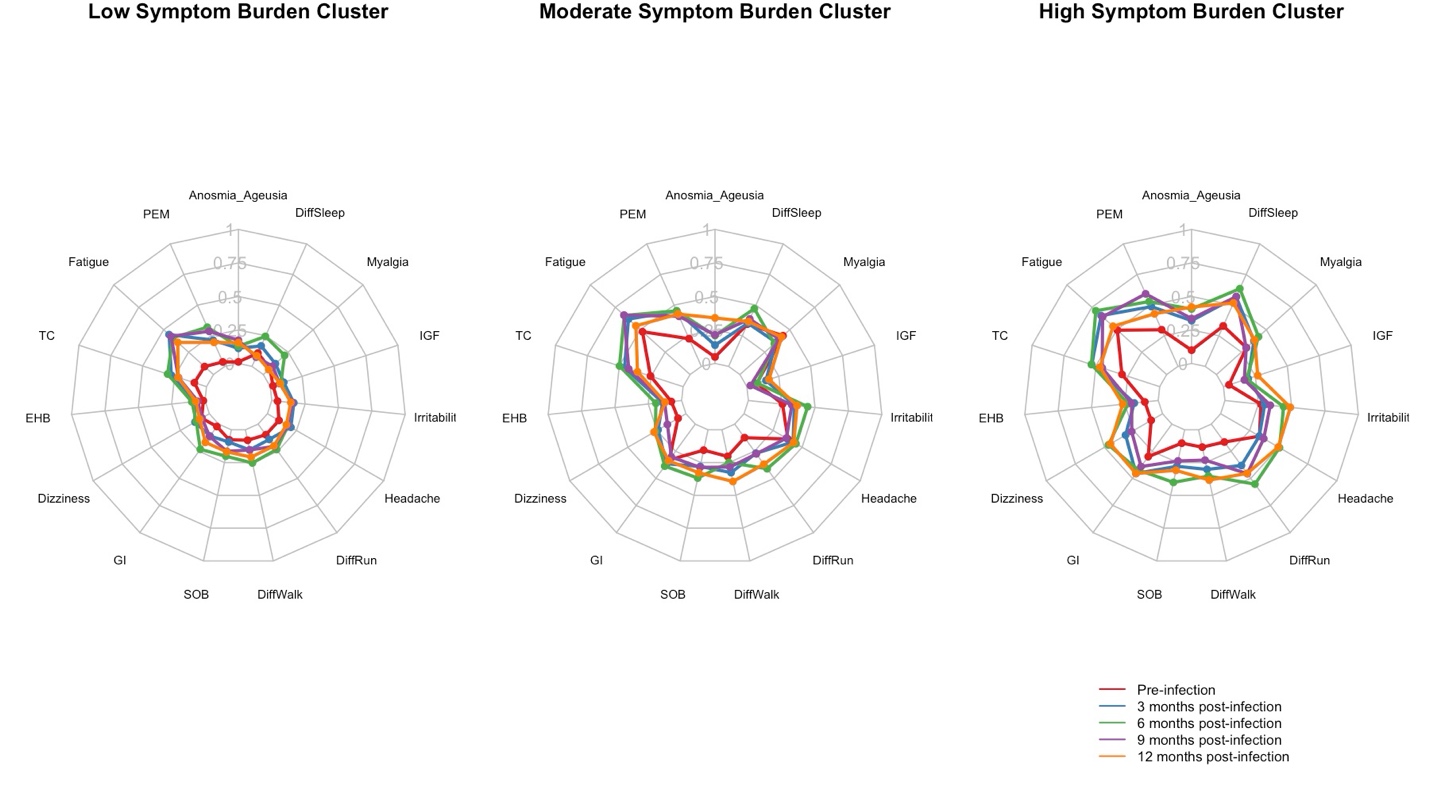

d. **Sensitivity analysis by excluding individuals who experienced a subsequent SARS-CoV-2 infection between 3 and 12 months after their index infection date**

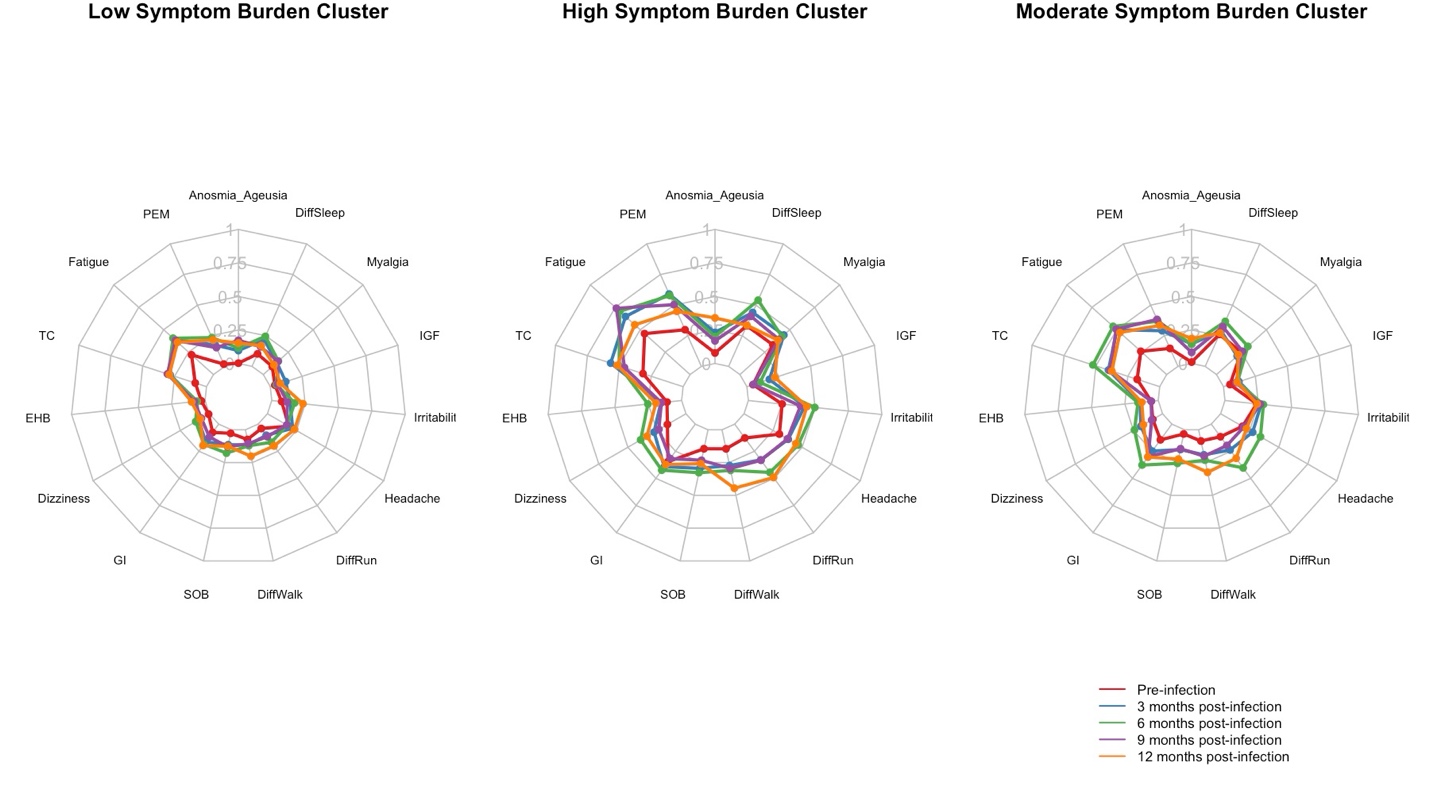

### eTable 1: Pooled Mean Proportion and 95% Confidence Intervals of Long COVID Symptoms Across Symptom Burden Clusters and Time Points, CHASING COVID Cohort Study (N = 511)

| **Cluster** | **Symptom** | **Pre-infection** | | **3-month post-infection** | | **6-month post-infection** | | **9-month post-infection** | | **12-month post-infection** | |
| --- | --- | --- | --- | --- | --- | --- | --- | --- | --- | --- | --- |
|  |  | **Mean** | **95% CI** | **Mean** | **95% CI** | **Mean** | **95% CI** | **Mean** | **95% CI** | **Mean** | **95% CI** |
| Cluster 1 | Difficulty running | 0.08 | 0.03,0.12 | 0.21 | 0.13,0.29 | 0.28 | 0.2,0.36 | 0.17 | 0.1,0.23 | 0.27 | 0.19,0.35 |
| Cluster 1 | Difficulty sleeping | 0.18 | 0.12,0.24 | 0.3 | 0.21,0.39 | 0.29 | 0.21,0.38 | 0.22 | 0.15,0.29 | 0.23 | 0.14,0.32 |
| Cluster 1 | Difficulty walking | 0.08 | 0.04,0.13 | 0.17 | 0.1,0.25 | 0.21 | 0.13,0.29 | 0.15 | 0.09,0.21 | 0.37 | 0.29,0.46 |
| Cluster 1 | Dizziness | 0.07 | 0.02,0.11 | 0.16 | 0.09,0.24 | 0.17 | 0.1,0.24 | 0.11 | 0.06,0.16 | 0.18 | 0.09,0.27 |
| Cluster 1 | Erratic Heartbeat | 0.03 | 0,0.07 | 0.11 | 0.05,0.18 | 0.15 | 0.06,0.23 | 0.07 | 0.02,0.12 | 0.15 | 0.04,0.26 |
| Cluster 1 | Fatigue | 0.17 | 0.11,0.23 | 0.46 | 0.37,0.56 | 0.44 | 0.35,0.52 | 0.43 | 0.35,0.51 | 0.42 | 0.33,0.52 |
| Cluster 1 | Gastrointestinal issues | 0.06 | 0.02,0.1 | 0.17 | 0.1,0.24 | 0.21 | 0.13,0.28 | 0.18 | 0.12,0.24 | 0.24 | 0.16,0.31 |
| Cluster 1 | Headache | 0.18 | 0.12,0.24 | 0.23 | 0.15,0.31 | 0.27 | 0.18,0.35 | 0.16 | 0.1,0.22 | 0.25 | 0.16,0.33 |
| Cluster 1 | Irritability | 0.1 | 0.05,0.15 | 0.23 | 0.15,0.31 | 0.22 | 0.14,0.29 | 0.19 | 0.12,0.26 | 0.26 | 0.17,0.35 |
| Cluster 1 | Low-grade fever | 0.06 | 0.02,0.11 | 0.02 | 0,0.05 | 0.13 | 0.06,0.2 | 0.07 | 0.02,0.11 | 0.15 | 0.07,0.22 |
| Cluster 1 | Loss of taste or smell | 0.03 | 0,0.06 | 0.11 | 0.04,0.17 | 0.15 | 0.07,0.24 | 0.1 | 0.05,0.15 | 0.26 | 0.16,0.35 |
| Cluster 1 | Post-exertional malaise | 0.08 | 0.03,0.12 | 0.31 | 0.22,0.4 | 0.28 | 0.2,0.36 | 0.23 | 0.16,0.29 | 0.32 | 0.23,0.41 |
| Cluster 1 | Myalgia | 0.12 | 0.06,0.17 | 0.16 | 0.09,0.23 | 0.2 | 0.13,0.28 | 0.14 | 0.08,0.2 | 0.15 | 0.07,0.23 |
| Cluster 1 | Shortness of breath | 0.1 | 0.06,0.15 | 0.14 | 0.08,0.21 | 0.22 | 0.14,0.3 | 0.15 | 0.09,0.21 | 0.21 | 0.13,0.29 |
| Cluster 1 | Trouble Concentrating | 0.1 | 0.05,0.15 | 0.31 | 0.23,0.39 | 0.39 | 0.31,0.48 | 0.27 | 0.2,0.34 | 0.33 | 0.24,0.42 |
| Cluster 2 | Difficulty running | 0.2 | 0.13,0.26 | 0.4 | 0.28,0.53 | 0.48 | 0.39,0.57 | 0.34 | 0.26,0.42 | 0.51 | 0.4,0.61 |
| Cluster 2 | Difficulty sleeping | 0.33 | 0.25,0.42 | 0.49 | 0.34,0.63 | 0.5 | 0.41,0.6 | 0.47 | 0.38,0.55 | 0.49 | 0.37,0.61 |
| Cluster 2 | Difficulty walking | 0.17 | 0.11,0.24 | 0.34 | 0.24,0.45 | 0.34 | 0.25,0.43 | 0.3 | 0.22,0.38 | 0.48 | 0.36,0.59 |
| Cluster 2 | Dizziness | 0.16 | 0.1,0.22 | 0.33 | 0.19,0.47 | 0.34 | 0.25,0.44 | 0.2 | 0.13,0.27 | 0.4 | 0.29,0.5 |
| Cluster 2 | Erratic Heartbeat | 0.13 | 0.07,0.19 | 0.21 | 0.1,0.33 | 0.25 | 0.15,0.34 | 0.12 | 0.06,0.19 | 0.28 | 0.17,0.39 |
| Cluster 2 | Fatigue | 0.51 | 0.43,0.6 | 0.74 | 0.63,0.85 | 0.69 | 0.6,0.78 | 0.63 | 0.55,0.71 | 0.57 | 0.44,0.7 |
| Cluster 2 | Gastrointestinal issues | 0.3 | 0.22,0.38 | 0.4 | 0.27,0.53 | 0.45 | 0.36,0.54 | 0.32 | 0.24,0.41 | 0.46 | 0.36,0.57 |
| Cluster 2 | Headache | 0.37 | 0.28,0.45 | 0.45 | 0.33,0.58 | 0.48 | 0.38,0.59 | 0.43 | 0.34,0.51 | 0.48 | 0.38,0.59 |
| Cluster 2 | Irritability | 0.34 | 0.26,0.42 | 0.39 | 0.26,0.51 | 0.54 | 0.44,0.63 | 0.35 | 0.27,0.44 | 0.46 | 0.34,0.58 |
| Cluster 2 | Low-grade fever | 0.05 | 0,0.09 | 0.06 | 0.01,0.12 | 0.14 | 0.06,0.21 | 0.1 | 0.05,0.16 | 0.27 | 0.17,0.36 |
| Cluster 2 | Loss of taste or smell | 0.07 | 0.02,0.11 | 0.26 | 0.14,0.38 | 0.28 | 0.17,0.39 | 0.19 | 0.12,0.26 | 0.37 | 0.26,0.47 |
| Cluster 2 | Post-exertional malaise | 0.3 | 0.22,0.38 | 0.52 | 0.37,0.66 | 0.56 | 0.47,0.66 | 0.46 | 0.37,0.54 | 0.53 | 0.42,0.65 |
| Cluster 2 | Myalgia | 0.38 | 0.3,0.47 | 0.48 | 0.37,0.6 | 0.46 | 0.36,0.55 | 0.44 | 0.35,0.53 | 0.49 | 0.38,0.6 |
| Cluster 2 | Shortness of breath | 0.15 | 0.09,0.21 | 0.31 | 0.19,0.42 | 0.32 | 0.23,0.41 | 0.27 | 0.19,0.35 | 0.33 | 0.22,0.43 |
| Cluster 2 | Trouble Concentrating | 0.35 | 0.27,0.43 | 0.55 | 0.43,0.67 | 0.61 | 0.51,0.7 | 0.46 | 0.38,0.55 | 0.53 | 0.41,0.65 |
| Cluster 3 | Difficulty running | 0.04 | 0.01,0.07 | 0.08 | 0.02,0.15 | 0.14 | 0.08,0.19 | 0.09 | 0.05,0.13 | 0.15 | 0.08,0.22 |
| Cluster 3 | Difficulty sleeping | 0.09 | 0.05,0.13 | 0.16 | 0.09,0.24 | 0.2 | 0.13,0.27 | 0.13 | 0.08,0.18 | 0.13 | 0.07,0.2 |
| Cluster 3 | Difficulty walking | 0.04 | 0.01,0.07 | 0.11 | 0.05,0.17 | 0.1 | 0.04,0.16 | 0.08 | 0.04,0.12 | 0.2 | 0.12,0.27 |
| Cluster 3 | Dizziness | 0.01 | 0,0.03 | 0.08 | 0.01,0.15 | 0.08 | 0.03,0.13 | 0.06 | 0.02,0.1 | 0.05 | 0,0.11 |
| Cluster 3 | Erratic Heartbeat | 0.02 | 0,0.03 | 0.08 | 0.02,0.14 | 0.05 | 0,0.1 | 0.03 | 0,0.06 | 0.09 | -0.02,0.21 |
| Cluster 3 | Fatigue | 0.11 | 0.06,0.15 | 0.3 | 0.22,0.39 | 0.31 | 0.24,0.38 | 0.26 | 0.19,0.32 | 0.24 | 0.16,0.31 |
| Cluster 3 | Gastrointestinal issues | 0.06 | 0.03,0.09 | 0.11 | 0.04,0.17 | 0.12 | 0.06,0.18 | 0.07 | 0.03,0.1 | 0.17 | 0.1,0.24 |
| Cluster 3 | Headache | 0.09 | 0.05,0.14 | 0.14 | 0.07,0.21 | 0.15 | 0.08,0.22 | 0.13 | 0.08,0.18 | 0.15 | 0.07,0.23 |
| Cluster 3 | Irritability | 0.04 | 0.01,0.07 | 0.12 | 0.05,0.2 | 0.14 | 0.08,0.2 | 0.1 | 0.05,0.14 | 0.16 | 0.09,0.23 |
| Cluster 3 | Low-grade fever | 0.02 | 0,0.04 | 0.01 | 0,0.03 | 0.05 | 0.01,0.09 | 0.04 | 0.01,0.07 | 0.04 | 0.01,0.08 |
| Cluster 3 | Loss of taste or smell | 0.03 | 0,0.05 | 0.1 | 0.04,0.17 | 0.08 | 0.02,0.14 | 0.06 | 0.02,0.09 | 0.15 | 0.08,0.21 |
| Cluster 3 | Post-exertional malaise | 0.02 | 0,0.04 | 0.15 | 0.07,0.23 | 0.11 | 0.06,0.16 | 0.12 | 0.07,0.17 | 0.17 | 0.09,0.25 |
| Cluster 3 | Myalgia | 0.02 | 0,0.05 | 0.12 | 0.05,0.19 | 0.09 | 0.03,0.14 | 0.05 | 0.02,0.08 | 0.07 | 0.01,0.12 |
| Cluster 3 | Shortness of breath | 0.02 | 0,0.05 | 0.07 | 0.02,0.12 | 0.1 | 0.05,0.15 | 0.06 | 0.02,0.1 | 0.08 | 0.03,0.12 |
| Cluster 3 | Trouble Concentrating | 0.05 | 0.02,0.08 | 0.19 | 0.12,0.27 | 0.19 | 0.12,0.26 | 0.16 | 0.11,0.22 | 0.19 | 0.12,0.26 |

### eTable 2: Optimal Number of Clusters Determined by NbClust Method Across 30 Imputed Datasets Using Multiple Clustering Indices

| Indices | Most common optimal number of clusters across 30 imputed datasets |
| --- | --- |
| Frey | 1 |
| McClain | 2 |
| C-index | 3 |
| Silhouette | 2 |
| Dunn | 10 |
| Hartigan | 3 |
| Ball and Hall | 3 |

### eTable 3: For each sensitivity analysis, adjusted Odds Ratios for Classification into High or Moderate Symptom Burden Clusters vs. Low Symptom Burden Cluster Among Participants with Long COVID, Based on K-Means Clustering, the CHASING COVID Cohort Study, December 2020 – December 2023

a. **Sensitivity analysis using Latent Class Analysis (LCA)**

|  | **High vs. low symptom burden cluster** | **Moderate vs. low symptom burden cluster** |
| --- | --- | --- |
|  | Adjusted Odds Ratios (95% CI) | Adjusted Odds Ratios (95% CI) |
| **Age** |  |  |
| ≤50 | ref | ref |
| >50 | **2.65 (1.58 – 4.36)** | **2.11 (1.28 – 3.84)** |
| **Gender** |  |  |
| Male | ref | Ref |
| Female | **1.98 (1.13 – 2.98)** | 1.05 (0.67 – 1.67) |
| Non-binary | 3.58 (0.96 – 8.01) | 0.58 (0.19 – 3.87) |
| **Number of comorbidities** |  |  |
| 0 | ref | ref |
| 1 | 1.07 (0.76 - 1.79) | 0.87 (0.26 - 1.87) |
| >=2 | **2.87 (1.76 – 7.07)** | 0.78 (0.41 – 2.18) |

b. **Sensitivity analysis defining long COVID cases as at least one symptom to be present at two distinct time points**

|  | **High vs. low symptom burden cluster** | **Moderate vs. low symptom burden cluster** |
| --- | --- | --- |
|  | Adjusted Odds Ratios (95% CI) | Adjusted Odds Ratios (95% CI) |
| **Age** |  |  |
| ≤50 | ref | ref |
| >50 | 1.12 (0.89 – 1.54) | 1.01 (0.80 – 1.22) |
| **Gender** |  |  |
| Male | ref | ref |
| Female | **1.74 (1.11 – 2.41)** | 1.78 (0.97 – 2.15) |
| Non-binary | 1.60 (0.87 – 4.35) | 1.77 (0.78 – 5.22) |
| **Number of comorbidities** |  |  |
| 0 | ref | Ref |
| 1 | 1.23 (0.84 – 2.37) | 1.17 (0.70 – 1.86) |
| >=2 | **2.57 (1.34 – 3.25)** | 1.58 (0.69 – 2.56) |

c. **Sensitivity analysis defining long COVID cases based on self-identified long COVID**

|  | **High vs. low symptom burden cluster** | **Moderate vs. low symptom burden cluster** |
| --- | --- | --- |
|  | Adjusted Odds Ratios (95% CI) | Adjusted Odds Ratios (95% CI) |
| **Age** |  |  |
| ≤50 | ref | ref |
| >50 | 1.64 (0.92 – 2.91) | 1.34 (0.70 – 2.57) |
| **Gender** |  |  |
| Male | ref | ref |
| Female | 1.32 (0.81 – 2.14) | 1.24 (0.73 – 2.10) |
| Non-binary | 3.07 (0.59 – 15.94) | 4.98 (1.05 – 23.50) |
| **Number of comorbidities** |  |  |
| 0 | ref | ref |
| 1 | **1.92 (1.10 – 3.34)** | 1.14 (0.64 – 2.06) |
| >=2 | **4.18 (2.19 – 7.98)** | **2.39 (1.20 – 4.76)** |

d. **Sensitivity analysis by excluding individuals who experienced a subsequent SARS-CoV-2 infection between 3 and 12 months after their index infection date**

|  | **High vs. low symptom burden cluster** | **Moderate vs. low symptom burden cluster** |
| --- | --- | --- |
|  | Adjusted Odds Ratios (95% CI) | Adjusted Odds Ratios (95% CI) |
| **Age** |  |  |
| ≤50 | ref | ref |
| >50 | **3.94 (2.14 – 7.26)** | **3.01 (1.72 – 5.26)** |
| **Gender** |  |  |
| Male | ref | ref |
| Female | 1.53 (0.89 – 2.62) | 1.36 (0.86 – 2.15) |
| Non-binary | 1.60 (0.38 – 6.69) | 2.07 (0.68 – 6.32) |
| **Number of comorbidities** |  |  |
| 0 | ref | ref |
| 1 | 1.00 (0.54 – 1.87) | 1.25 (0.75 – 2.06) |
| >=2 | **2.87 (1.41 – 5.83)** | 1.72 (0.89 – 3.32) |
